# Is Lower Limb Movement Enough? Quantifying Overground Arm Swing Kinematics and Coordination to Assess Ageing Decline in Older Adults

**DOI:** 10.64898/2026.08.03.26359452

**Authors:** Kai Zhe Tan, Sai G.S. Pai, Yong Kuk Kim, Angela Frautschi, Michelle Gwerder, Kok Yang Tan, Vanessa Jean Wen Koh, Deepak K. Ravi, William R. Taylor, Rahul Malhotra, Angelique Wei-Ming Chan, David B. Matchar, Navrag B. Singh

**Affiliations:** Future Health Technologies Programme, Singapore-ETH Centre, Singapore, Singapore.; Institute for Biomechanics, ETH Zurich, Zurich, Switzerland.; Programme in Health Services Research & Population Health (HSRPH), Duke-NUS Medical School, Singapore.; Centre for Ageing Research & Education (CARE), Duke-NUS Medical School, Singapore.

**Author notes:** Contributing authors.

**Keywords:** Arm swing, wearable sensors, ageing, fall risk

## Abstract

Traditional clinical gait assessments focus on lower-limb kinematics and overall walking speed, often overlooking the upper-limb dynamics and inter-limb coordination that matter for real-world ambulation. Measuring these movements outside the laboratory is difficult, and this study presents and validates a wearable sensor algorithm to quantify arm swing kinematics and arm-leg coordination (Phase Locking Value, PLV) during overground walking in the home. Validated against optical motion capture, the algorithm detected swing events reliably and with negligible temporal bias. In home-based gait recordings from nearly 1500 community-dwelling older adults, arm-leg coordination was the strongest arm swing predictor of rhythmic gait stability once walking speed was accounted for. Exploratory factor analysis separated upper-limb function into distinct “coordination and stability” and “pace and capacity” axes, identifying arm swing as an independent dimension of the gait profile. Analysis of dynamic resilience during turns showed that frail older adults have slower recovery of arm-leg coordination, pointing to a loss of motor automaticity. Across a 30-year age span, arm swing amplitude declined with age while arm–leg coordination did not change detectably. In a separate laboratory cohort, coordination was also reduced in Parkinson’s disease, indicating that the measure responds to neurological impairment as well as to frailty. This algorithm offers a scalable way to assess upper-limb gait dynamics in daily life. Shifting the clinical focus from walking speed alone to full-body movement may help detect instability early.

## 1 Introduction

The evolution of bipedalism freed the upper limbs but left the body less stable dynamically. The rhythmic, bi-phasic swing of the arms is one adaptation that helps to overcome this instability. Holding the arms still during walking increases metabolic cost by about 12% and raises the peak twisting moment between the foot and the ground by roughly 63% [1]. Oscillating in a bi-phasic manner, arm swing works as a counterweight that regulates angular momentum and stabilises the centre of mass [2]. Controlling lateral balance is itself metabolically costly: providing external lateral stabilisation reduces both step width variability and the energy cost of walking, in older as well as young adults [3]. Restricting the arms does not by itself reduce the stability of steady-state walking, but it does slow the return to the normal gait pattern after a perturbation, which indicates that it is the recovery movements of the arms that contribute to overall stability [4]. Actively increasing arm swing also improves local dynamic stability, particularly in the medial-lateral direction [5], which matters for preventing injurious falls. Arm swing may therefore be a useful training target: in healthy older adults, brief seated training of arm-swing rhythm with a wearable robot increased stride length and walking speed and reduced the proportion of double-limb support [6], and perturbation-based balance training reduces fall rates among older adults, including those who are frail [7].

Arm swing and gait kinematics have typically been measured with optical motion capture systems in the laboratory. However, these setups do not necessarily capture what happens in real-world walking – the turns, pauses, and unexpected perturbations that can lead to falls [8, 9]. While wearable inertial measurement units (IMUs) allow continuous, overground kinematic assessment of arm swing [10, 11], current IMU-based methods derive swing amplitude from angular velocity without establishing the direction of travel. Arm swing therefore cannot be expressed relative to the plane of progression, and changes in walking direction are not accounted for. An algorithm that estimates and corrects sensor orientation (heading) continuously, and without relying on a magnetometer, is therefore required for valid free-living assessment of arm swing and inter-limb coordination.

Although arm swing changes naturally with age, specific deviations in amplitude, symmetry, and coordination may signal underlying neuromuscular conditions [12, 13]. Reduced arm swing amplitude, increased asymmetry and reduced bilateral coordination are candidate prodromal markers for neurological disorders such as Parkinson’s disease, and have been detected in at-risk individuals before other major motor deficits appear [14, 15]. More broadly, wearable and smartphone-based gait measures are being developed as sensitive digital markers of disability progression in other neurological conditions such as multiple sclerosis [16]. Ageing affects the protective use of the arms as well: older adults accelerate their arms more slowly than young adults when responding to a slip, which reduces the stabilising contribution of the arm reaction and leaves them more vulnerable to falls [17]. Despite this diagnostic value, the translation of upper-limb kinematics into standard geriatric assessment is limited by a lack of data collected outside the laboratory. Quantitative arm swing parameters – swing variability and inter-limb coordination indices – together with a description of how they vary with age and frailty status are therefore needed. With them, these continuous metrics could function as non-invasive biomarkers to detect early neuromotor change and track the progression of fall risk in daily life. Consumer-grade smartwatches, already worn by millions of older adults, offer a ready deployment platform for such algorithms, making population-level arm swing assessment possible without disrupting standard clinical protocols.

We develop a wearable-sensor algorithm (Figure 1) that extracts continuous arm swing kinematics and discrete swing events during overground walking, validated against optical motion capture as a gold standard. We then use it to explore the role of upper-limb movement in overall gait using Exploratory Factor Analysis and quantify step-by-step recovery of inter-limb coordination after turns using Phase Locking Value (PLV). Finally, we describe how arm swing parameters vary across a 30-year age span in older adults. This framework and its reference values allow upper-limb movement, gait stability and coordination to be assessed in real-world walking.

**Fig. 1:**
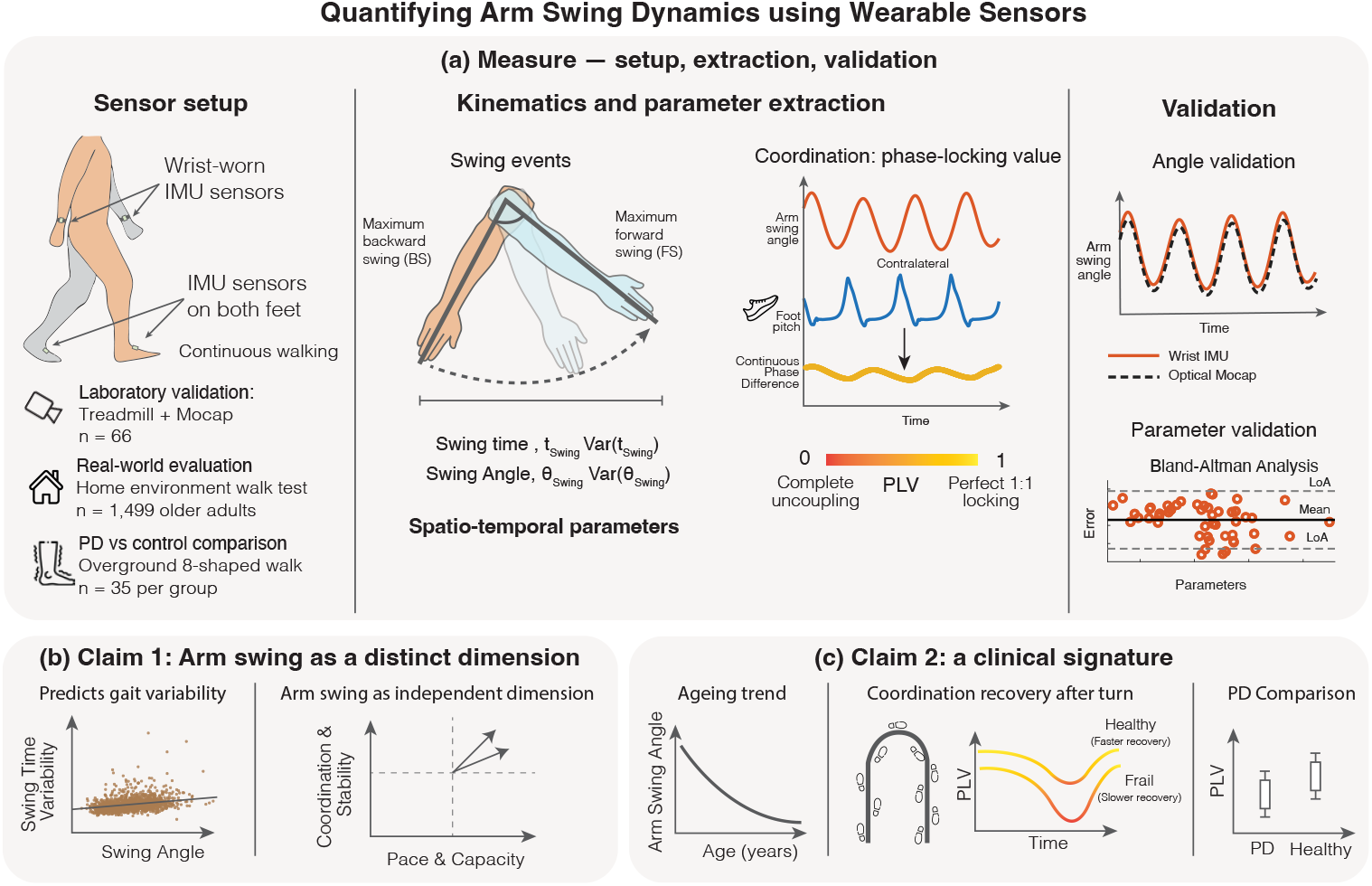
Schematic overview of the wearable sensor arm swing algorithm and data analysis framework:(a) Sensor setup, kinematic extraction, and validation against optical motion capture. (b) Arm swing predicts gait variability and separates from lower-limb measures in factor analysis. (c) Clinical signature: amplitude declines with age, coordination recovery after turns is slower with frailty, and coordination is reduced in Parkinson’s disease.

## 2 Results

### 2.1 A wearable algorithm measures arm swing and inter-limb coordination accurately

We benchmarked the algorithm against optical motion capture on three criteria: continuous trajectory reconstruction, temporal accuracy of swing event detection, and agreement of the derived parameters (Figure 2; full results in Appendix A, Tables A1, A2 and A3). Agreement is reported throughout as the Intraclass Correlation Coefficient in its single-measure, absolute-agreement form, ICC(A,1). By estimating the orientation dynamically with adaptive PCA, the proposed method removed the orientation drift of existing algorithms (Fan et al. [10] and Warmerdam et al. [11]), reducing the Root Mean Square Error (RMSE) from 8.3 4.9° and 8.8 4.1° to 3.6 2.0°, and tracked the reference closely (Coefficient of Multiple Correlation (CMC)=0.93; Pearson’s r=0.94). Maximal Forward and Backward Swing events were identified with over 99% sensitivity and few false detections (F1-score = 0.99), with timing errors of 2.4 8.6 ms and 6.3 ± 14.4 ms respectively. Arm Swing Time showed no systematic bias (Bias = 0.00 s; ICC(A,1) = 0.998); because this metric is the interval between consecutive backward swing events, any constant detection offset cancels in the difference, so the event timings above are the more informative test of temporal accuracy. Agreement was also high for the variability measures (Arm Swing Time Variability ICC(A,1) = 0.966; Arm Swing Amplitude Variability ICC(A,1) = 0.963), with biases of 0.11 and 0.67 percentage points of the coefficient of variation, while Arm Swing Amplitude was underestimated by 3.5*^◦^* at a reference mean of 43.2*^◦^* (relative error 9.2%; ICC(A,1) = 0.936), as expected from a single distal sensor that does not capture dynamic elbow flexion. Amplitude Asymmetry showed the weakest agreement (ICC(A,1) = 0.848, mean absolute error 2.55 percentage points), reflecting the propagation of bilateral amplitude error into a ratio measure.

**Fig. 2:**
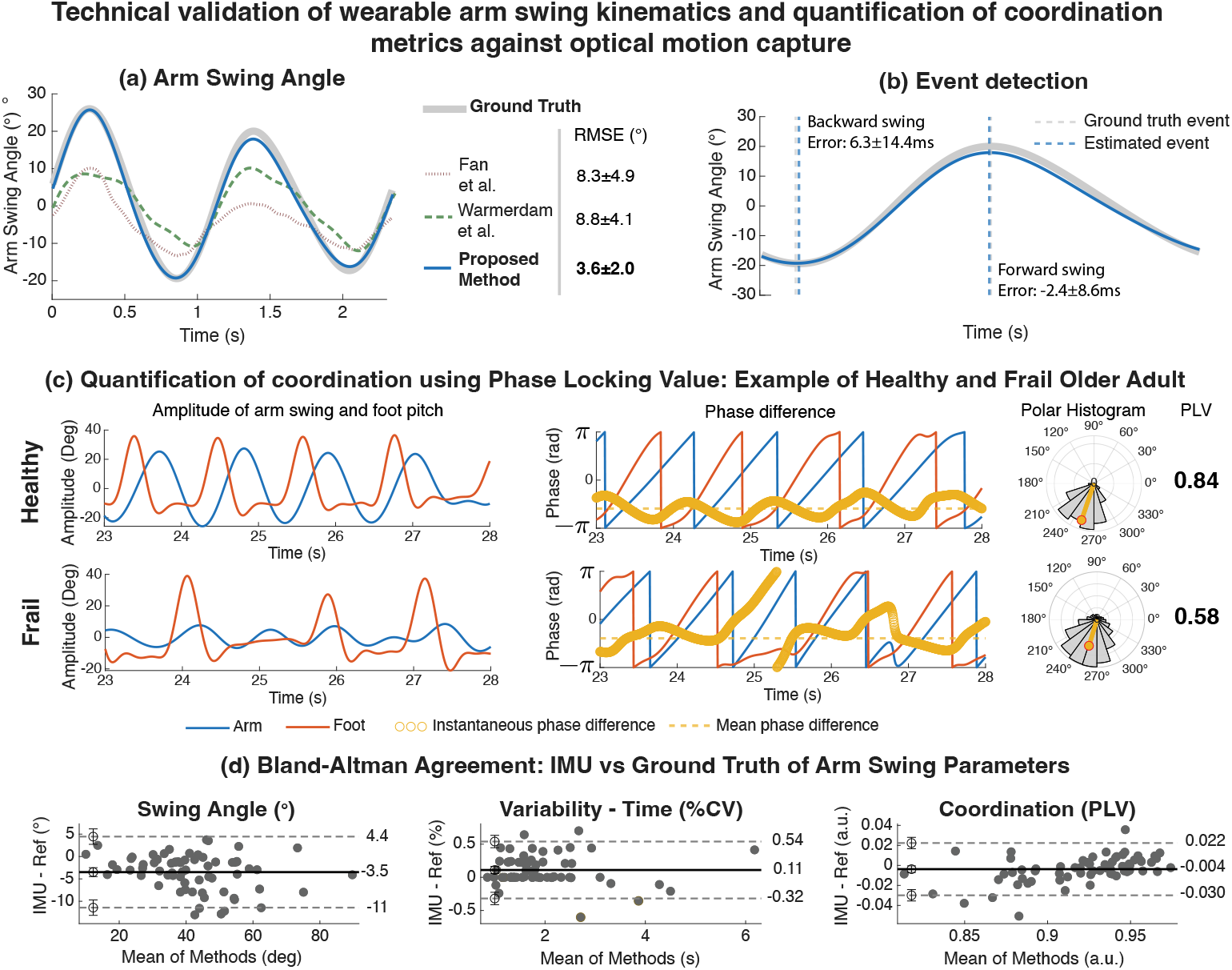
Technical validation of wearable arm swing kinematics and coordination metrics: (a) Representative trajectories show that the proposed orientation-correction algorithm (blue) eliminates the orientation drift typical of existing methods (Fan et al. 2025, Warmerdam et al. 2020), closely tracking the optical motion capture gold standard (grey line). (b) Temporal validation shows detection of gait events with high accuracy (F1-score = 0.99) and minimal lag. (c) The arm-leg Phase Locking Value (PLV) captures distinct coordination patterns: healthy gait (top) maintains stable 1:1 phase locking, whereas frail gait (bottom) exhibits wandering phase differences. PLV = 1 represents perfect locking, PLV = 0 represents complete randomness. (d) Bland-Altman analyses for Arm Swing Amplitude, Arm Swing Time Variability, and PLV. Agreement with optical motion capture is high for the variability and coordination measures.

Coordination between the contralateral upper and lower limbs (e.g. left wrist and right foot) was quantified using the PLV, which combines the temporal and spatial aspects of coordination into a single metric derived from the continuous arm swing angle and foot pitch. Similarly, agreement with the optical reference was high (ICC(A,1) = 0.929; Bias =-0.004; relative error 1.1%), and the metric separated the stable phase-locking of a healthy participant from the dispersed phase control of a frail one (Figure 2c). **From this agreement, the minimal detectable change for one participant is 0.027 PLV units at 95% confidence, which is the resolution at which an individual’s coordination can be tracked over repeated assessments. Group mean differences smaller than this remain detectable, because the standard error of a group mean scales with** 1*/ n*. Together, these findings show that paired wrist-and foot-worn sensors can capture the small fluctuations of inter-limb coordination during real-world walking. With the measurement in hand, we turned to overground walking in older adults, and to what arm-leg coordination shows about locomotor health.

### 2.2 Arm-leg coordination is a distinct dimension of gait, independent of walking speed

We first asked whether arm-leg coordination is a real, separable part of gait, or just a byproduct of how fast and how far the limbs move. Two questions follow: does coordination matter for gait stability beyond what walking speed explains, and does it stand apart statistically from lower-limb and arm-amplitude measures?

#### 2.2.1 Arm-leg coordination predicts gait variability independent of walking speed

Arm swing metrics were mapped against two domains of gait performance: pace (gait speed, Figure 3 Top Row) and lower limb gait variability (swing time variability adjusted for speed, Figure 3 Bottom Row), the latter representing rhythmic stability. All arm swing metrics were significantly associated with walking speed (Pearson’s correlation coefficient, *r* = 0.22 0.52), but their relationship with gait variability changed after controlling for the confounding effect of speed. Arm Swing Amplitude explained little of the variance in gait variability (Partial *R*^2^=0.056). Similarly, temporal variability of the arm swing (Partial *R*^2^=0.078) and amplitude variability (Partial *R*^2^ =0.094) showed only moderate associations.

**Fig. 3:**
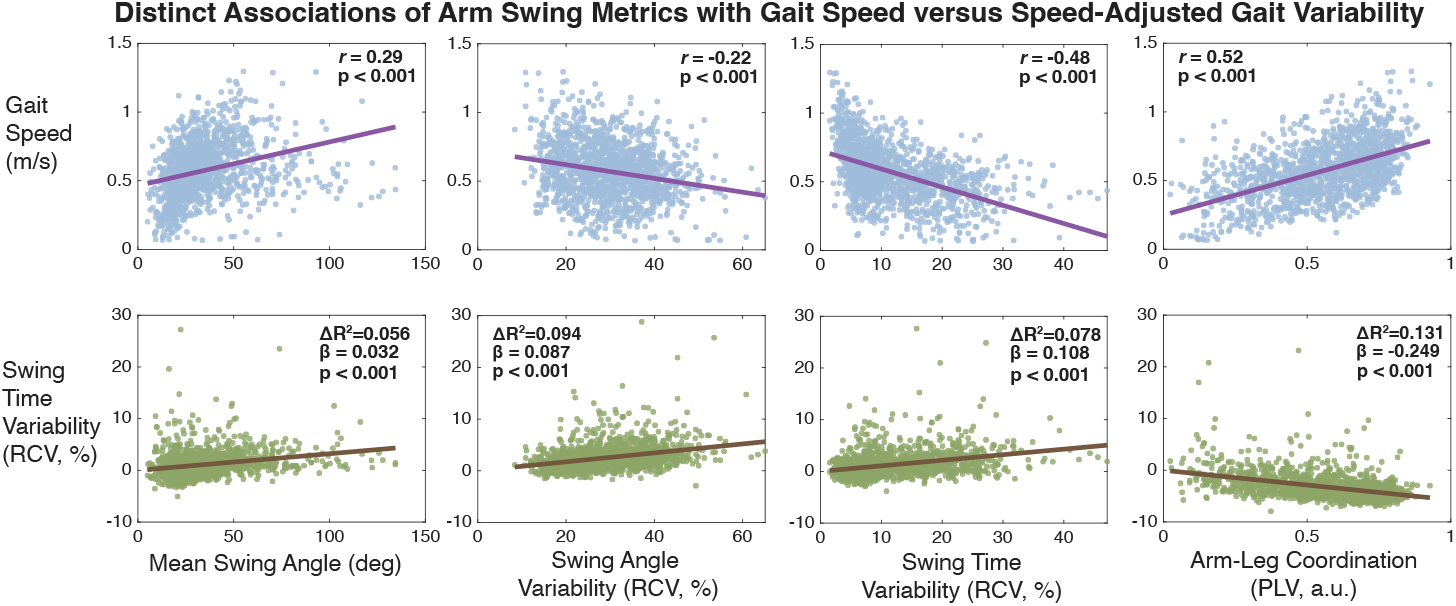
Distinct associations of arm swing metrics with gait speed and speed-adjusted gait variability. Top row: Relationships between arm swing parameters and gait speed (walking pace). Pearson’s correlation coefficients (*r*) indicate that arm swing amplitude and variability metrics are coupled with walking speed (purple lines). Bottom row: Relationships between arm swing parameters and lower limb Swing Time Variability (gait rhythmic stability) after adjusting for gait speed and age. Partial residual plots (brown lines) display the unique variance explained (Partial *R*^2^) and the regression slope (*β*). Higher Arm Swing Amplitude is associated with increased variability (positive *β*, indicating noise), whereas higher coordination (PLV) is associated with decreased variability (negative *β*, indicating stability). All associations displayed are statistically significant (*p <* 0.001).

Arm-leg PLV was the strongest arm swing predictor of gait variability. Even after adjusting for walking speed, it explained 13.1% of the residual variance (Partial *R*^2^ = 0.131, *p <* 0.001), more than any unilateral arm metric. Dominance analysis showed a clear hierarchy of predictors (Table 1; total model *R*^2^ = 0.463). Gait speed was the primary determinant of gait variability, accounting for 44.8% of the model’s total explained variance, followed by arm-leg PLV at 26.0%. Arm Swing Amplitude contributed only 1.6%. This indicates that the temporal synchronisation of the limbs is a stronger predictor of rhythmic stability than the kinematics of the arm swing itself, and that some of the instability observed in frail gait is associated with degraded inter-limb coordination rather than slower walking speed alone.

**Table 1:**
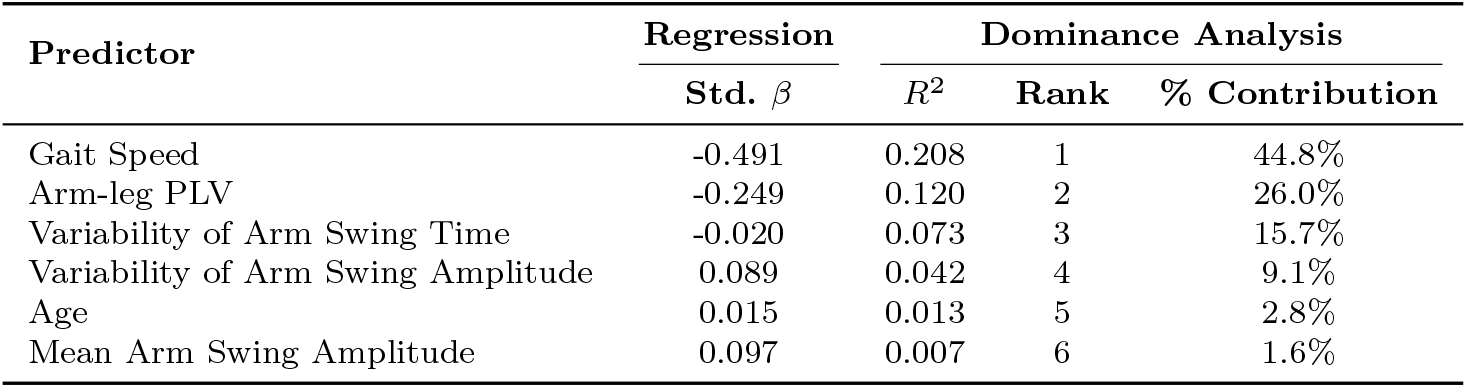
Dominance Analysis of Arm Swing Metrics, separating the unique contribution of each parameter from the shared variance.

| Predictor | Regression | Dominance Analysis |  |  |
| --- | --- | --- | --- | --- |
| | Std. $\beta$ | $R^2$ | Rank | % Contribution |
| Gait Speed | -0.491 | 0.208 | 1 | 44.8% |
| Arm-leg PLV | -0.249 | 0.120 | 2 | 26.0% |
| Variability of Arm Swing Time | -0.020 | 0.073 | 3 | 15.7% |
| Variability of Arm Swing Amplitude | 0.089 | 0.042 | 4 | 9.1% |
| Age | 0.015 | 0.013 | 5 | 2.8% |
| Mean Arm Swing Amplitude | 0.097 | 0.007 | 6 | 1.6% |

#### 2.2.2 Arm swing kinematics form an independent dimension of the overall gait profile

Regression analysis showed coordination to be an independent predictor of stability, but this result leaves unexplained whether arm swing parameters represent a distinct domain of locomotor control or whether they are simply a byproduct of lower limb movement. To test this, we ran an Exploratory Factor Analysis on the full gait parameter set (Table 2) and a focused analysis of arm swing metrics (Table 3).

**Table 2:** Rotated Factor Matrix for Gait and Arm Swing Parameters: Bold values indicate *|*loadings*| >* 0.5. *|*Loadings*| <* 0.2 are omitted. F1=Rhythm, F2=Rhythmic Stability, F3=Arm Swing Variability, F4=Pace, F5=Spatial Stability, F6=Lower-limb Swing Timing. F6 is not the primary loading for any variable. The highest loadings of F6 are Avg Swing Time (0.683) and Cadence (*−*0.449), listed under F1.

| Gait variable | F1 | F2 | F3 | F4 | F5 | F6 |
| --- | --- | --- | --- | --- | --- | --- |
| **Rhythm (F1)** |  |  |  |  |  |  |
| Avg Stride Time | **0.953** |  |  |  |  |  |
| Avg Step Time | **0.954** |  |  |  |  |  |
| Avg Stance Time | **0.951** |  |  | -0.204 |  |  |
| Avg DLS Time | **0.923** |  |  | -0.263 |  |  |
| Cadence | **-0.809** |  |  | 0.227 |  | -0.449 |
| Avg Swing Time | **0.517** |  |  |  |  | **0.683** |
| **Rhythmic Stability (F2)** |  |  |  |  |  |  |
| Var Step Time | 0.218 | **0.925** |  |  | 0.212 |  |
| Var Swing Time | 0.258 | **0.821** | 0.215 | -0.225 | 0.204 |  |
| Var DLS Time |  | **0.754** |  |  | 0.312 |  |
| Var Stance Time |  | **0.660** | 0.446 |  | 0.434 | 0.243 |
| Var Stride Time |  | **0.641** | 0.458 |  | 0.408 |  |
| Asymmetry |  | **0.586** |  |  |  |  |
| **Arm Swing Variability (F3)** |  |  |  |  |  |  |
| Var Arm Swing Time | 0.290 |  | **0.792** |  |  |  |
| Arm-leg PLV | -0.353 |  | **-0.769** |  | -0.229 |  |
| Var Arm Swing Amplitude |  |  | **0.753** |  | 0.201 |  |
| **Pace (F4)** |  |  |  |  |  |  |
| Avg Stride Length | -0.314 |  |  | **0.899** |  |  |
| Avg Step Length (L) |  |  |  | **0.776** | -0.236 | 0.214 |
| Avg Gait Speed | **-0.523** |  |  | **0.743** |  | -0.270 |
| Arm Swing Amplitude | -0.223 |  | 0.295 | 0.291 |  |  |
| **Spatial Stability (F5)** |  |  |  |  |  |  |
| Var Gait Speed |  | 0.349 | 0.329 | -0.209 | **0.832** |  |
| Var Stride Length |  | 0.232 | 0.247 | -0.242 | **0.812** |  |
| Var Step Length (L) | 0.255 | 0.401 |  | -0.201 | **0.523** |  |
| **Variance Expl.** | 24.8% | 17.7% | 12.6% | 11.7% | 11.4% | 4.7% |
| **Cumulative Var** | 24.8% | 42.5% | 55.1% | 66.8% | 78.2% | 82.9% |

**Table 3:** Rotated Factor Matrix for Upper Limb Parameters. This focused factor analysis identifies the two factors of arm swing behaviour. Bold values indicate *|*loadings*| >* 0.5. *|*Loadings*| <* 0.2 are omitted. Factor 1 represents Coordination and Stability, while Factor 2 represents Pace and Capacity of arm swing.

| Variable | Factor 1 | Factor 2 |
| --- | --- | --- |
| **Coordination & Stability** |  |  |
| Arm-leg PLV | **0.897** |  |
| Variability of Arm Swing Time | **-0.875** |  |
| Variability of Arm Swing Amplitude | **-0.718** | 0.345 |
| Arm Swing Amplitude Asymmetry |  | -0.216 |
| **Pace & Capacity** |  |  |
| Mean Arm Swing Time |  | **-0.696** |
| Mean Arm Swing Amplitude |  | 0.474 |
| **Cross-Loading** |  |  |
| Gait Speed | **0.643** | **0.717** |
| **Variance Explained** | 36.0% | 20.1% |
| **Cumulative Variance** | 36.0% | 56.2% |

For the whole structure of gait (Table 2), the parameters are separated into six orthogonal dimensions explaining 82.9% of the total variance (Parallel Analysis identified six dimensions as optimum). Several arm swing metrics, consisting of spatio-temporal variability and arm-leg PLV formed an independent factor (Factor 3), distinguishing from lower-limb temporal rhythmicity (Factor 1) and step-to-step variability (Factor 2). Factor 3, representing Arm Swing Variability, accounts for 12.6% of the total variance in the gait parameter space, marking it as a substantial, independent locomotor domain.

The focused analysis on the upper limb parameters (Table 3) separated two underlying groups: Factor 1 (“coordination and stability”) loaded heavily on arm-leg PLV (0.897) and Arm Swing Time Variability (*−*0.875), capturing the consistency of the movement pattern, while Factor 2 (“pace and capacity”) loaded on Mean Arm Swing Time ( −0.696), capturing the magnitude of the movement. Mean Arm Swing Amplitude loaded on the same factor at 0.474, below the 0.5 threshold used here. Gait Speed was shared, cross-loading on both factors (0.643 on coordination and stability, 0.717 on pace and capacity). Arm Swing Amplitude Asymmetry loaded weakly on both ( −0.216 at most), indicating that side-to-side imbalance varies largely independently of the two axes. Overall, this suggests that while walking speed reflects the ability to maintain healthy spatiotemporal strides, the control needed for precise arm-leg coordination (Factor 1) is different from the physical capacity driving arm swing magnitude and pace (Factor 2).

### 2.3 Coordination shows no age-related change but is reduced with frailty

We then asked whether coordination carries clinical meaning. If it reflects a neural control process rather than general physical capacity, it should hold steady across older age yet respond once the neuromotor system that maintains it is challenged or damaged. We looked for this pattern across three settings – older age, frailty, and Parkinson’s disease.

#### 2.3.1 Arm swing amplitude declines with age while coordination does not

To characterise how arm swing varies across older age, we modelled trajectories for men and women aged 60 to 90 years using Generalised Additive Models for Location, Scale and Shape (GAMLSS), in the 1,476 participants of the home-based cohort who fell within this age range (847 women, 629 men; 23 were older than 90). This analysis showed a clear dissociation between the physical magnitude of movement and the neural control of rhythm.

Arm Swing Amplitude declined steadily with age in both sexes (Figure 4, left panels), reflecting a natural loss of locomotor capacity or musculoskeletal flexibility. The decline was clear in both sexes when tested against a model with no age term (women: likelihood-ratio *χ*^2^(1) = 32.3, *p <* 0.001; men: *χ*^2^(1) = 15.2, *p <* 0.001), amounting to a fall in fitted median amplitude of 12.2*^◦^*in women (95% CI 8.0 to 16.3*^◦^*) and 8.4*^◦^*in men (95% CI 4.2 to 12.5*^◦^*) across the 30-year span.

**Fig. 4:**
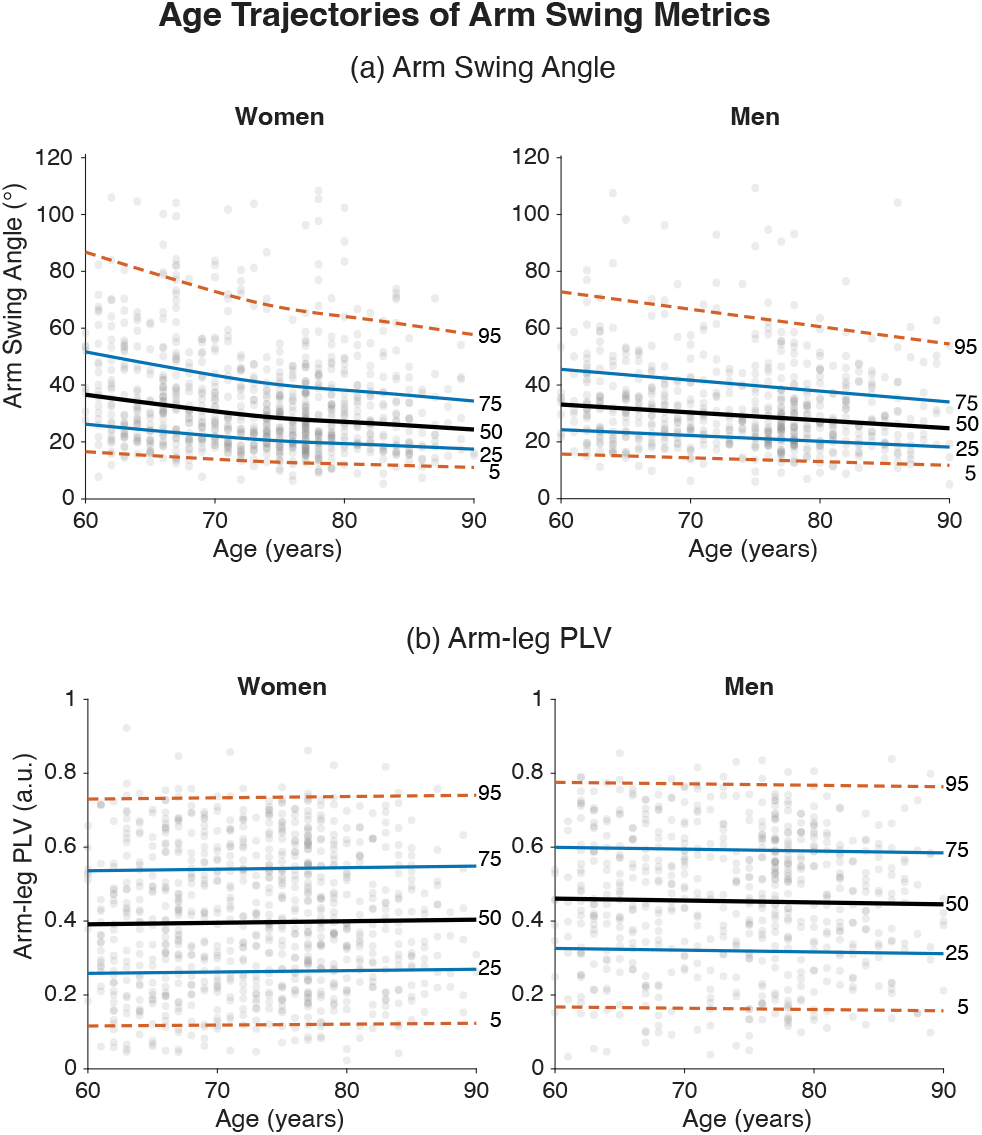
Age trajectories of Arm Swing Amplitude and Inter-limb Coordination: GAMLSS-fitted centiles for home-based Arm Swing Amplitude (left) and arm-leg PLV (right), in 1,476 participants aged 60 to 90 years. Arm Swing Amplitude declined progressively with advancing age in both sexes, consistent with general losses in locomotor capacity. The bands for PLV are flat across the 30-year span, and the age term did not improve model fit in either sex (both *p >* 0.6).

In contrast, arm–leg PLV showed no detectable trend with age (Figure 4, right panels): the fitted centile bands were flat across the age range in both sexes. Testing the age term directly, neither a linear age effect (women: *χ*^2^(1) = 0.18, *p* = 0.67; men: *χ*^2^(1) = 0.23, *p* = 0.63) nor a penalised smooth (women: *χ*^2^(2.1) = 4.87, *p* = 0.10; men: *χ*^2^(1.4) = 1.07, *p* = 0.41) improved on a model without age. The fitted median PLV changed by +0.013 in women and 0.015 in men between ages 60 and 90, both well inside the 0.027 minimal detectable change computed in the validation cohort. Arm swing amplitude and inter-limb coordination therefore behave differently across this age range. Whether the stability of the coupling reflects preserved control or simply an insensitive measure cannot be settled from these trajectories alone, so we next asked whether the same measure responds to an everyday challenge and to disease.

#### 2.3.2 Recovery of coordination is slower for frail compared to healthy older adults

We next used arm-leg PLV to ask how quickly coordination is restored after a turn. Turns act as natural perturbations that disrupt steady-state walking and are a known challenge for older adults, contributing to falls [18, 19]. Of 18,614 turns detected across the home-based recordings, 1,334 participants had enough straight-line walking around a turn to give a coordination value at every step in the window (116 non-frail, 1,030 pre-frail, 188 frail; median 7 turns each, IQR 4 to 13). All groups showed the same “V-shaped” trajectory (Figure 5a): PLV dropped during the turn and recovered over the following steps. Coordination before the turn was already lower with increasing frailty (0.731, 0.680 and 0.637 in non-frail, pre-frail and frail adults; both differences *p <* 0.001), and the recovery that followed was slower.

**Fig. 5:**
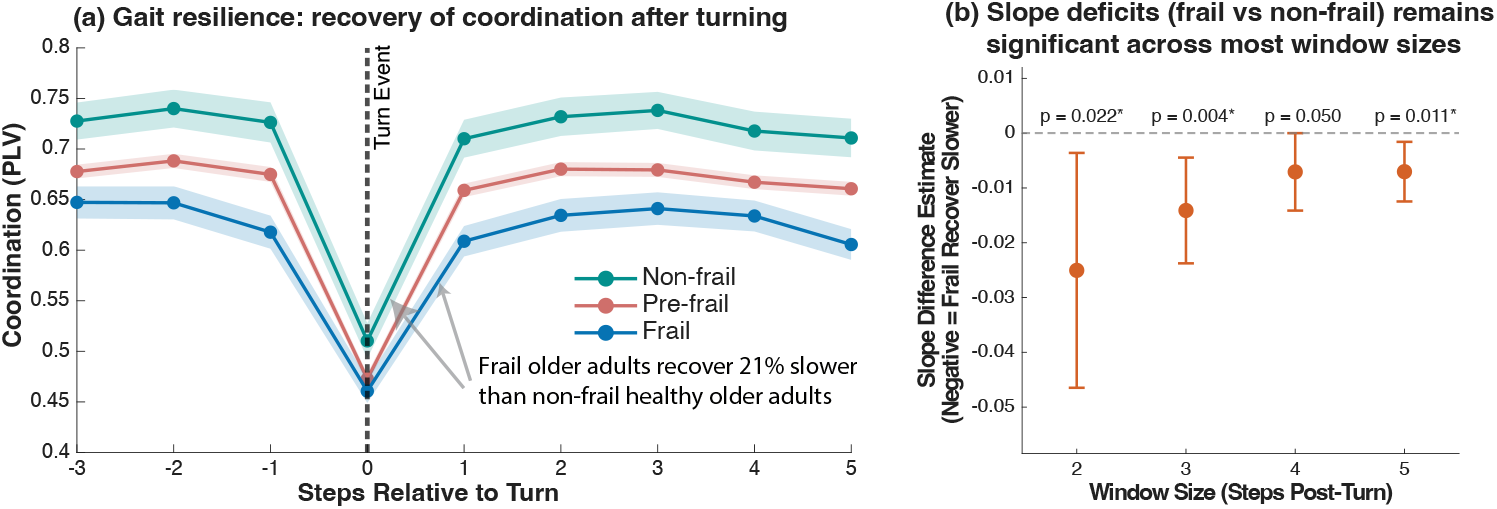
Dynamic resilience and recovery of arm-leg PLV following a turn. (a) Trajectories of arm-leg PLV before and after a turn event (Step 0), stratified by clinical frailty phenotype. While all groups experience an expected mechanical drop in phase-locking during the turn, recovery slows progressively with frailty status: pre-frail adults regain baseline coordination 11% more slowly and frail adults 21% more slowly than non-frail older adults. Estimates are from a mixed model weighted by the number of turns each participant contributed. Shaded regions represent 95% confidence interval. (b) Recovery slope deficits (frail vs. non-frail) across varying post-turn windows (2–5 steps). Estimates are negative at every window length and reach significance at three of the four, indicating a consistent direction of effect.

Non-frail older adults recovered at 0.069 PLV units per step (95% CI 0.062 to 0.075). Pre-frail adults recovered 11.2% more slowly (*β*_interaction_ = *−*0.008, 95% CI 0.015 to 0.001, *p* = 0.034) and frail adults 20.6% more slowly (*β*_interaction_ = − 0.014, 95% CI 0.024 to 0.005, *p* = 0.004). Recovery therefore slowed step by step with frailty status, which suggests a progressive loss of the reserve needed to restore stability after an everyday perturbation. The deficit was consistent across post-turn windows of 2 to 5 steps (Figure 5b). Sensitivity analyses that weighted, adjusted for, or restricted on the number of turns each participant contributed gave the same result, with one exception: when the sample was limited to the 542 participants with at least ten turns, the estimate was smaller and no longer significant (Table A4).

#### 2.3.3 Coordination is reduced in Parkinson’s disease

If rhythmic arm-leg coordination does not change across older age, is it affected by neurological disease? We tested this in patients with Parkinson’s disease. This comparison used a laboratory dataset recorded with optical motion capture rather than the wearable system (previously reported by Mei et al. [20]; Amprimo et al. [21]), and is therefore a test of the PLV construct itself rather than of the wearable algorithm. As the two groups were recruited independently and the controls were on average 6.9 years older, we first age-matched them one-to-one, giving 35 pairs with no residual age difference (both groups 62.5 years). We compared Arm Swing Amplitude, Asymmetry and PLV between these 35 matched controls and 35 individuals with PD recorded before Deep Brain Stimulation (DBS) surgery.

Arm Swing Amplitude was significantly reduced in PD relative to healthy controls (Hedges’ *g* = −0.60, 95% CI [ 1.08, 0.12]), about a 22% reduction. Arm Swing Asymmetry was substantially greater in the PD group (*g* = 0.75 [0.27, 1.24]), reflecting greater side-to-side imbalance. PLV was significantly lower in PD (*g* = − 0.73 [ 1.21, 0.24]), indicating degraded bilateral arm-leg coordination. A linear mixed-effect model gave group differences in the same direction and of comparable magnitude (Table 4). The PD–control difference in PLV of 0.040 units exceeds the 0.027 minimal detectable change of the wearable system, though we note that this comparison was made with optical markers on a fixed path, so it does not by itself show that the wearable would resolve the same difference in home conditions. Taken together, these results show that arm-leg coordination is separable from pace and lower-limb movement, stays steady across healthy age, and is sensitive to both frailty and neurological disease.

**Table 4:** Arm swing comparison between Parkinson’s Disease and healthy control in the age-matched sample, measured with optical motion capture. LMM *β* is reported in the units of each measure.

| Measure | Control<br>$M$ (SD) | PD<br>$M$ (SD) | Hedges’ $g$ [95% CI] | LMM $\beta$<br>(SE) | $p_{\text{LMM}}$ |
| --- | --- | --- | --- | --- | --- |
| Amplitude (°) | 24.1 (7.9) | 18.7 (9.6) | $-0.60$ [$-1.08, -0.12$] | $-5.38$ (2.07) | .009 |
| Asymmetry | 10.3 (5.1) | 16.7 (10.9) | $0.75$ [$0.27, 1.24$] | $6.47$ (2.01) | .001 |
| PLV | 0.860 (0.017) | 0.820 (0.075) | $-0.73$ [$-1.21, -0.24$] | $-0.040$ (0.013) | .002 |

*Note.* $M$ = participant mean; SD = between-participant standard deviation. Hedges’ $g$ is from independent-samples $t$ -tests on participant means in the age-matched sample (Control $n = 35$ , PD $n = 35$ ); all three differences were significant ( $p \leq .013$ ), and the confidence intervals exclude zero. LMM $\beta$ = fixed-effect estimate from the segment-level model with Control as reference; SE = standard error. Positive $g$ for Asymmetry indicates higher (worse) values in PD; negative $g$ for Angle and PLV indicates lower (worse) values in PD.

## 3 Discussion

Using a wearable-sensor algorithm validated against optical motion capture, this study quantifies home-based arm swing parameters and arm–leg coordination in older adults, and shows their value as markers of locomotor health beyond walking speed. The algorithm quantified arm swing parameters, their variability, and inter-limb coordination through the Phase Locking Value (PLV) in settings where earlier methods could not. Applied to home-based recordings, this framework produced several findings about gait of older adults:

1. PLV was validated as a measure for coordination, and explained gait variability well after adjusting for gait speed;
2. Exploratory factor analysis showed that arm swing kinematics form a distinct dimension of overall gait;
3. Arm Swing Amplitude declined with age, whereas arm-leg coordination did not change detectably across the same range;
4. Recovery of coordination following a turn slowed progressively with frailty status, from non-frail through pre-frail to frail older adults;
5. Parkinson’s Disease (PD) patients showed worse arm swing coordination, smaller swing amplitude and greater asymmetry than healthy controls.

Together, these findings challenge the view that arm swing is a passive, mechanical byproduct of lower limb movement. Gait speed was the single largest contributor to gait variability, accounting for 44.8% of the explained variance, but PLV contributed to gait stability independent of walking speed, explaining a meaningful share of the remaining variance (partial *R*^2^=0.131). This indicates that inter-limb phase locking plays an active role in managing step-to-step fluctuations. The factor analysis supported this finding: a separate axis for arm swing variability, on which arm swing spatio-temporal variability and inter-limb coordination loaded highly. Taken together, the results highlight the value of directly measuring arm swing to capture a fuller picture of gait. The arm-swing-specific factor analysis separated two axes: “pace and capacity” (Arm Swing Amplitude and gait speed) and “coordination and stability” (arm-leg PLV and swing variability). This distinction matters clinically: an older adult who walks quickly with a large Arm Swing Amplitude (high capacity) may still lack the neural coordination to stabilise that movement against everyday perturbations (low stability) – and walking fast without rhythmic stability could increase their fall risk. Quantifying PLV during walking in the home allows identification of coordination deficits, that speed-based assessments such as the 4-metre walk test miss, enabling clinicians to identify these hidden subtle deficits and intervene in time.

Our findings show that arm-leg coordination (PLV) predicts real-world rhythmic stability, consistent with evidence linking poor inter-limb coordination to higher fall risk in older adults [22, 23]. The lower baseline PLV in frail older adults, together with their slower post-turn recovery, indicates that these individuals possess limited functional reserve and reduced automaticity in locomotor control [24]. The inter-limb coupling that PLV captures reflects a locomotor control system that humans share with other mammals. In animal models, each limb is controlled by its own spinal central pattern generator, and these networks are coordinated by long propriospinal pathways linking the cervical and lumbar cord, together with somatosensory feedback and descending supraspinal pathways [25]. In humans, rhythmic arm movement during walking arises from both passive biomechanical linkages and active neural commands, and the arm–leg coordination it produces is thought to draw on these same conserved circuits [25, 26]. This layered organisation may help explain the pattern we observed: the basic arm–leg coupling appears stable and largely automatic, showing no change across the age range we modelled (Section 2.3.1), yet dependent on intact neural circuitry – hence the reduced PLV in individuals with PD (Section 2.3.3), where dopaminergic degeneration specifically disrupts the basal ganglia organisation that drives rhythmic locomotor automaticity [27, 28]. Adjusting that coupling to meet a more complex demand, such as restoring coordination after a turn, relies on more flexible control that is vulnerable in frail individuals (Section 2.3.2). Turning also demands prefrontal executive control beyond that of steady-state walking [29, 30]. Hence, a frail functional reserve, already challenged during straight line walking, is quickly overwhelmed [24, 31]. This cognitive-motor overload is a plausible explanation for the delayed inter-limb coordination recovery we observed, and illustrates how a loss of automatic neural control may leave frail individuals vulnerable to falls during daily life.

Clinically, separating locomotor capacity from coordination control matters for accurate geriatric assessment. Age-related declines in Arm Swing Amplitude likely reflect normal physiological changes with age, such as sarcopenia, joint stiffness, or more cautious gait [32]. Arm–leg coordination behaved differently: PLV showed no age trend across the 30-year span we modelled. Earlier reports of age-related change in inter-limb coordination compare young with older adults and largely concern coupling between the legs [22]; in the treadmill validation cohort, young (n=41, 23.7 years) and older adults (n=25, 68.0 years) differed in PLV by only 0.008 units, well below the 0.027 minimal detectable change and not statistically significant (*p* = 0.36). A drop in arm–leg coordination in an older adult is therefore more likely to signal a neurological deficit than ordinary physical decline. The clear reduction in PLV observed in individuals with PD – a large effect size, derived from wrist and foot kinematics alone – points to the diagnostic sensitivity of these coordination metrics and their potential as screening tools for neuromotor dysfunction across the clinical spectrum. Given that arm swing can be captured continuously and unobtrusively from wearable sensors in free-living conditions, inter-limb coordination indices could offer a scalable, low-burden complement to assessments such as the MDS-UPDRS or Timed-Up-and-Go test. Future work should test whether inter-limb coordination is a modifiable risk factor, for example, whether rhythmic auditory stimulation, dual-tasking, or complex agility training can restore PLV and preserve sensorimotor plasticity in pre-frail populations [31].

This study has three main limitations. First, the design is cross-sectional, so we cannot describe how arm swing changes within a person over time, and the minimal clinically important difference for these metrics remains to be established; both require longitudinal follow-up. Second, the algorithm was validated on treadmill walking. Sensor heading drifts on a treadmill as it does overground, and the adaptive-PCA step corrected that drift here, so the same correction should extend to the larger heading changes that turning produces. However, showing this directly would nonetheless require overground recordings against optical motion capture, which this study does not include. Third, the home recordings were supervised and confined to a single session, and participants chose their own route, so the number of turns available differed between them. We weighted each participant by their turn count and repeated the analysis across window lengths, turn-count thresholds and model specifications (Table A4). The deficit held in every setting except the most restrictive turn-count threshold. Recording over several days would supply more turns per person and allow how much a person walks to be separated from how well they recover.

## 4 Conclusion

In summary, this study measured overground arm swing kinematics with a wearable sensor algorithm, yielding metrics for locomotor capacity, variability, and coordination. While traditional clinical evaluations heavily prioritise lower-limb movements and overall walking speed, arm swing measures added information about gait beyond walking speed and lower-limb parameters in older adults. By providing a validated method to measure arm swing under real-world conditions, this work offers a new instrument for evaluating coordination and rhythmic stability, potentially targeting underlying locomotor deficits before falls occur.

## 5 Methods

### 5.1 Data Collection and Processing

#### 5.1.1 Study participants and data collection protocol

This study used data from three different cohorts: (a) a lab-based study with IMU and motion capture system for technical validation of arm swing parameters, (b) a homebased overground walking study for evaluation of arm swings in older adults and (c) a Deep Brain Stimulation (DBS) study cohort that consists of Parkinson’s Disease (PD) patients and healthy controls. All participants provided written informed consent, and the respective study protocols were approved by the local institutional review boards, adhering to the principles of the Declaration of Helsinki. Ethical approval was granted by the Cantonal Ethics Commission Zurich for the lab-based treadmill cohort (EK-2021-N-145) and by the National University of Singapore Institutional Review Board for the home-based cohort (NUS-IRB-2021-168); approval for the DBS cohort is stated in Section 5.1.4. Table 5 summarises the demographics of all three cohorts.

**Table 5:** Demographics of the lab-based treadmill cohort, the home-based overground walking cohort and the DBS Parkinson’s disease cohort.

| Cohort | Group | n | Age (years)<br>M (SD) | Range<br>(years) | Female<br>n (%) |
| --- | --- | --- | --- | --- | --- |
| Lab-based treadmill | Young | 41 | 23.7 (2.3) | 19–29 | 24 (58.5%) |
|  | Older | 25 | 68.0 (4.5) | 60–77 | 16 (64.0%) |
| Home-based overground | — | 1499 | 73.8 (7.6) | 60–99 | 861 (57.4%) |
| DBS, age-matched | Control | 35 | 62.5 (10.3) | 40–77 | n.r. |
|  | PD | 35 | 62.5 (10.1) | 41–77 | 7 (20.0%) |

*Note.* The home-based row describes the 1,499 participants with complete gait, demographic and frailty data; the number contributing to each analysis is given in the relevant Results section and in Section 5.1.3. The DBS rows describe the age-matched sample used in Section 2.3.3, drawn from 51 controls and 54 patients; the two groups were matched one-to-one on age and do not differ ( $p = 1.00$ ). n.r. = not recorded; sex was not available for the DBS control group.

#### 5.1.2 Lab-based treadmill cohort

Data were collected from 100 participants recruited from the local community in Zürich, Switzerland, in two age groups: 50 young adults (23.8 ± 2.3 years) and 50 older adults (68.8 ± 5.7 years). Inclusion criteria were individuals with no known neurological disorders, movement dysfunction, or fall history. Each participant walked continuously on a treadmill for six minutes at a constant, self-selected comfortable speed. Synchronised wearable and optical recordings suitable for validation were obtained from 66 of them, and these 66 form the validation sample reported here: 41 young adults (23.7 ± 2.3 years, range 19 to 29; 24 women, 58.5%) and 25 older adults (68.0 ± 4.5 years, range 60 to 77; 16 women, 64.0%).

For validation ground truth, gait kinematics were recorded at 100 Hz using a 10-camera optical motion capture system (Vicon Nexus v2.3/2.8.2, Oxford Metrics, UK) with 61 reflective markers. Wearable gait data were recorded using Inertial Measurement Unit (IMU) sensors (ZurichMove, Zürich, Switzerland [33, 34]). Sensors were placed on both feet, on the pelvis and on both wrists. Each IMU captured triaxial acceleration and tri-axial angular velocity with a dynamic range of ±16 g and ±2000°/s, respectively, sampled at 200 Hz and time-synchronised across all sensors.

#### 5.1.3 Home-based overground walking cohort

Data were drawn from the Targeted Assessment and Recruitment of Geriatrics for Effective fall prevention Treatments (TARGET) cohort study for assessment of falls and fracture risk in community-dwelling older adults in Singapore. The study is a cross-sectional investigation with stratified random sampling of older adults across Singapore. The study was conducted in the participant’s own home environment. Participants were instructed to walk continuously for at least five minutes, at home in their preferred setting. They were free to turn as needed, and a study interviewer closely followed behind for safety. IMU data were collected with the same sensor model, placement (both wrists, both feet, pelvis) and sampling configuration as the lab-based cohort.

1,499 participants with wrist sensor data had complete demographic and frailty data and form the home-based cohort described in Table 5 (73.8 ± 7.6 years, range 60 to 99; 861 women, 57.4%). The number contributing to each analysis differs according to the variables it requires: 1,497 for the speed-adjusted regressions, dominance analysis and factor analyses; 1,476 for the normative GAMLSS models, which additionally require the participant to fall within the modelled age range of 60 to 90 years; and 1,334 for the turn-recovery analysis, which requires a complete peri-turn coordination profile (Section 2.3.2).

#### 5.1.4 DBS PD study cohort

We used a previously collected dataset introduced by Mei et al. [20] and Amprimo et al. [21], Gait kinematics were available before STN-DBS electrode implantation, while ON-medication, for 54 individuals diagnosed with PD, and for 51 healthy controls evaluated once. The groups were recruited independently and were not age-matched by design: the PD patients were 59.7 ± 10.5 years old (range 36 to 77) and the controls 66.6 ± 10.8 years (range 40 to 85), a difference of 6.9 years (*p* = 0.001). The comparison therefore used an age-matched subsample of 35 pairs, implemented in the MatchIt package for R [35]. Matching was one-to-one without replacement, both groups averaged 62.5 years.

Within that subsample the PD patients had a mean age at symptom onset of 53.4 ± 9.5 years, a disease duration of 9.1 ± 4.4 years, a Hoehn and Yahr stage of 2.1 ± 0.5. Inclusion criteria for the PD group were a diagnosis of idiopathic Parkinson’s disease, age between 18 and 90 years, designation for STN-DBS implantation at University Hospital Zurich, and the ability to walk independently and continuously for 10 minutes. Participants in the lab-based treadmill cohort and this cohort were separate samples with no overlap. Participants walked barefoot at a self-selected speed for 10 continuous minutes, without assistance, along an 8-shaped path around two markers positioned 10 m apart. Only straight-path segments were recorded. Gait kinematics were recorded at 100 Hz using a 10-camera optical motion capture system (Vicon Nexus v2.3/2.8.2, Oxford Metrics, UK) with 61 reflective markers. All arm swing and coordination parameters for this cohort were derived directly from the marker trajectories, using the same definitions applied to the wearable data. Ethical approval was granted by the Cantonal Ethics Commission Zurich (Protocol no. 2015-00141), and all participants provided written informed consent.

#### 5.1.5 Wearable Data Processing and Gait Parameter Extraction

The lab-based treadmill and home-based overground cohorts were both recorded with wearable IMUs and processed as follows. For each walk, key gait events (heel-strikes and toe-offs) were then extracted from the foot sensors across the full recording using a previously validated event-detection algorithm from the tri-axial accelerometer and gyroscope signals [34]. As non-stationary movements can confound variability metrics, we also applied validated turn-and pause-detection algorithms [33] and discarded pauses longer than 5 seconds and turns exceeding 45*^◦^*. This restricted the analysis to continuous straight-line walking, so that the resulting metrics reflected near steady-state gait variability in the home-based overground setting.

A set of spatio-temporal parameters was then derived per gait cycle from these events. The temporal parameters – stride, swing, stance, step, and double-limb-support times – followed directly from the event timings [33]. The spatial parameters, stride length and gait speed, were obtained by strapdown integration of the foot accelerometer signals, with sensor orientation estimated using the Versatile Quaternion-based Filter (VQF) [36]. Variability was quantified as the Robust Coefficient of Variation:

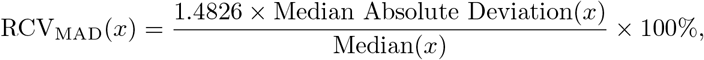

where the constant 1.4826 rescales the median absolute deviation so that it estimates the standard deviation for normally distributed data, making RCV_MAD_ directly comparable in magnitude to the conventional coefficient of variation [37].

### 5.2 Arm Swing Quantification with Wearable Sensors

To quantify the behaviour of arm swing using wearable IMU sensors, we extracted continuous arm swing trajectories, discrete gait events, and spatiotemporal parameters from the wrist-worn IMU data. This process mirrors standard lower limb gait analysis algorithms [33], where continuous kinematics are broken down into cycles (analogous to heel strikes in lower-limb analysis) to enable cycle-to-cycle variability assessment [23].

#### 5.2.1 Continuous Drift-Free Arm Swing Trajectories Estimation from IMU Sensors

The Arm Swing Trajectory (*θ*) is the overall angular displacement of the forearm segment projected onto the instantaneous plane of progression. Biomechanically, this projection isolates the anterior-posterior contribution of the limb to locomotor rhythm, omitting non-sagittal components such as arm abduction or circumduction. To quantify Arm Swing Angle in real-world environments without reliance on magnetometers (which are susceptible to ferromagnetic interference), we developed a drift-correction algorithm based on adaptive Principal Component Analysis (PCA). First, raw sensor orientation quaternions (*q*) were converted into a global rotation matrix to extract the forearm unit vector, **v**_forearm_. Since the gravity vector (*g* = [0, 0, 1]) provides a stable vertical reference, the primary challenge was correcting the drift in the horizontal orientation (yaw).

We assumed that during walking, the dominant axis of arm motion occurs in the sagittal plane (anterior-posterior direction). These are the steps to estimate this heading vector dynamically:

1. **Horizontal projection**: The vertical component of the forearm vector was subtracted to isolate the horizontal trajectory.
2. **Sliding window PCA**: We applied PCA to the horizontal forearm trajectory over a sliding window of 2.0 seconds (approx. 2 gait cycles). The first principal component (eigenvector corresponding to the largest eigenvalue) was identified as the instantaneous axis of arm swing.
3. **Heading alignment**: To prevent 180° flipping (i.e. forward to backward), the eigenvector was continuously aligned with the previous heading estimate using a dot-product continuity check.
4. **Signal smoothing**: The resulting heading vector was low-pass filtered (Butterworth filter, 4th order, cut-off 0.1 Hz) to remove step-to-step oscillations while preserving the trajectory of the walking path.

Finally, the arm swing angle *θ*(*t*) was evaluated by projecting the forearm vector onto the sagittal plane defined by the estimated heading and the gravity vector (as shown in Figure 1). The angle was computed as the deviation between the projected vector and the vertical, where positive values indicate forward swing and negative values indicate backward swing, as stipulated by the following equation:

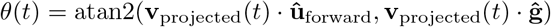

where **v**_projected_ is the projected forearm vector, **ĝ** is the gravity vector defining the vertical axis, and **û**_forward_ is the drift-corrected heading vector defining the forward axis. This formulation ensures a continuous signed angle where 0° corresponds to the neutral vertical position, positive values denote forward swing, and negative values denote backward swing.

#### 5.2.2 Determining arm swing events

Gait events were automatically detected from the continuous arm swing angle signal using a peak detection algorithm (findpeaks, Signal Processing Toolbox, MATLAB R2025b [38]) validated against optical motion capture.

- Maximal Forward Swing (FS): Defined as the local maxima of the angle signal (peak flexion).
- Maximal Backward Swing (BS): Defined as the local minima of the angle signal (peak extension).

For each gait cycle *i*, the Arm Swing Amplitude was calculated as the range of motion. Arm Swing Time was calculated as the temporal difference between consecutive backward swing events. To ensure analysis of steady-state gait, turns and non-walking periods were automatically excluded based on the angular velocity of the pelvis-worn IMU.

The FS event was used as the primary temporal point for the arm swing cycle – analogous to the Heel Strike in lower-limb gait analysis. Just as the heel strike marks the initiation of the stance phase and sets the cadence of the legs, the FS marks the distinct maximal point of the arm’s pendular movement, providing a marker for extracting cycle-to-cycle timing and variability.

#### 5.2.3 Derivation of Spatio-temporal Arm Swing Parameters

Based on these events, the following spatiotemporal arm swing parameters were calculated for each gait cycle *i*:

- Arm Swing Amplitude, *θ*_Swing_: Quantifying the total range of motion within a cycle, calculated as the difference of angles between FS and BS *i*:

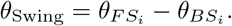

- Arm Swing Time, *t*_swing_: The temporal duration of one complete arm cycle, defined as the time interval between consecutive BS events

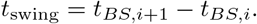

- Arm Swing Time Variability and Arm Swing Amplitude Variability, Var(*t*_swing_) and Var(*θ*_Swing_): The Robust Coefficient of Variation, RCV_MAD_ as defined above, of the arm swing time and of the arm swing amplitude respectively [37], serving as metrics of rhythmic stability.
- Arm Swing Asymmetry: The symmetry angle of bilateral swing amplitudes 45° *−* arctan(*X_R_/X_L_*) */*90° *×* 100, quantifying the difference in amplitude between the left and right arm swing. [39]

#### 5.2.4 Quantifying upper and lower limb coordination using Phase Locking Value

To quantify the stability of inter-limb coordination, we sought a single-valued metric that captures the strength of synchronisation between the contralateral arm and leg. Phase-based measures of limb coupling have precedent in this setting: using wrist-worn accelerometers, Huang et al. [15] showed that the variability of the instantaneous relative phase between the two arms was elevated in Parkinson’s disease, and that this loss of coordination was distinguishable from the amplitude asymmetry more commonly reported. That work addressed bilateral arm–arm coupling; the arm–leg coupling of interest here requires a metric with the same phase-based logic but applied across the upper and lower limb. Although the Phase Coordination Index (PCI) [40] is widely used in gait analysis, it was developed to characterise the anti-phased timing of left–right stepping (bilateral lower-limb coordination) and is therefore not intended to capture the 1:1 frequency locking between the upper and lower limbs that is of interest here. Continuous Relative Phase (CRP) offers a more detailed, time-continuous phase relationship, but the choice of summary statistic matters [41]. Linear summaries such as mean absolute relative phase or deviation phase can be distorted by phase-wrapping, because phase is a circular quantity confined to [ − *π, π*] and thus jumps discontinuously at the boundary. For example, a set of phase differences clustered near a half-cycle boundary (e.g. +178° and 178°, just 2° apart across the wrap) represents a constant coordination state, yet a linear average returns approximately 0° erroneously implying in-phase coupling. To overcome these limitations and obtain a direct quantification of coordination, we used the Phase Locking Value (PLV) [42]. Rather than averaging the phase-difference values, PLV represents each phase difference as a unit vector and computes the magnitude of their mean resultant vector. As this operation has no boundary discontinuity, it is inherently insensitive to phase wrapping; in the example above, the samples across the wrapping boundary point in nearly the same direction and yield a high PLV, correctly reflecting stable coupling. PLV is mathematically equivalent to the circular concentration of the CRP distribution, and returns a normalised value between 0 and 1 that is independent of movement amplitude or the specific phase lag angle, where 1 denotes perfectly consistent coupling and 0 denotes random phase drift.

First, the instantaneous phase *ϕ*(*t*) of the arm swing trajectory and the contralateral foot pitch were extracted using the Hilbert Transform. This transformation converts the real-valued kinematic signal *x*(*t*) into a complex analytical signal *Z*(*t*):

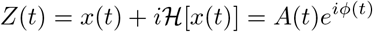

where *H* is the Hilbert transform, *A*(*t*) is the instantaneous amplitude envelope, and *ϕ*(*t*) is the instantaneous phase within [*−π, π*]. The continuous relative phase difference between arm swing and foot pitch was then calculated as Δ*ϕ*(*t*) = *ϕ*_arm swing_(*t*) *− ϕ*_foot pitch_(*t*).

Then, the PLV was computed as the length of the mean resultant vector of the relative phase distribution over the duration of the walking bout (over *N* data samples):

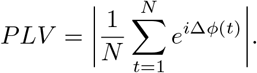

The resulting value ranges from 0 to 1, providing a normalised index of synchronization consistency. PLV = 1 indicates perfect 1:1 frequency locking with a constant phase lag (strict coupling), while PLV = 0 indicates complete uncoupling or random drift. In the context of upper and lower limbs coordination, a high PLV reflects a gait controller that maintains synchronisation despite environmental noise. Conversely, a reduced PLV indicates poor arm-leg coupling, characterizing a frailty-associated degradation in the precise timing of inter-limb coordination.

### 5.3 Statistical analysis

#### 5.3.1 Validation and Comparison

The proposed wearable system was assessed against the optical motion capture across three dimensions: continuous waveform fidelity, event detection precision, and parameter agreement. Continuous arm swing trajectories were evaluated using Root Mean Square Error (RMSE), Pearson’s correlation coefficient (r), and the Coefficient of Multiple Correlation (CMC) [43] to quantify waveform similarity and tracking accuracy. Discrete gait event detection (Maximal Backward and Forward Swing) was assessed using F1-scores, Sensitivity, and Positive Predictive Value (PPV) [34] to determine temporal precision. Finally, the agreement of derived spatiotemporal and coordination parameters (e.g., Amplitude, Swing Time, PLV) was quantified using the single-measure, absolute-agreement Intraclass Correlation Coefficient, ICC(A,1) [44], and Bland-Altman analysis [45] to visualise systematic bias and limits of agreement (LoA). Left and right sides were averaged within a participant before computing agreement statistics. From these, the standard error of measurement was obtained as 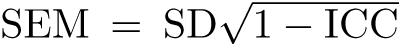 and the minimal detectable change at 95% confidence as 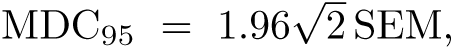 giving the smallest change in an individual that can be distinguished from measurement error.

#### 5.3.2 Differential Association of Arm Swing Metrics

Baseline associations between arm swing metrics and gait stability were visualised using scatter plots with fitted regression lines. To isolate speed-independent contributions, we calculated the Partial Coefficient of Determination (Partial *R*^2^) for each arm swing metric, adjusting for walking speed. This quantified the proportion of residual variance in gait variability explained by arm kinematics after removing the confounding effect of locomotor speed.

To establish a hierarchy of predictors, we constructed a multivariate linear model including all potential determinants (Speed, PLV, Amplitude, Arm Swing Time Variability, Amplitude Variability, and Age). We then conducted a Dominance Analysis [46] to decompose the model’s total explained variance (*R*^2^). The General Dominance Weight (average marginal contribution across all subset models) was calculated for each predictor, allowing us to rank variables by their relative importance in preserving gait stability independent of model specification order.

#### 5.3.3 Exploratory Factor Analysis

Two exploratory factor analyses were run on the 1,499 participants of the homebased cohort, both using varimax (orthogonal) rotation in R [47]. The full-model analysis entered 22 spatio-temporal gait parameters (means and variability of stride, stance, swing, step and double-limb-support time; stride and step length; gait speed; cadence; asymmetry) together with the four arm swing measures, and extracted six factors by minimum residual. The focused analysis entered the seven upper-limb and pace measures listed in Table 3 and extracted two factors by the same method. The number of factors was set by parallel analysis with 100 iterations. Loadings above 0.5 in absolute value are treated as substantial and those below 0.2 are omitted from the tables.

#### 5.3.4 Quantification of Dynamic Resilience - Turn Recovery Analysis

To assess the dynamic resilience, we analysed the recovery of arm-leg coordination following a turning manoeuvre. Turning represents a natural perturbation that transiently disrupts steady-state walking, requiring the system to actively restore the continuous phase-locking pattern.

To process the data for walking windows selection, turning events were automatically detected using the yaw angular velocity of the pelvis-worn IMU exceeding 90°*/s*, an established threshold that corresponds to deliberate body rotation (a rate that produces a 90° change in walking direction if sustained for approximately one second) and excludes the smaller rotations associated with postural sway during straight-line walking. This turn detection is separate from, and serves the opposite purpose to, the turn exclusion described in Section 5: the steady-state analyses discard segments containing turns above 45° so that variability metrics reflect straight-line walking, whereas the present analysis deliberately selects turns to study recovery from them. We extracted walking windows consisting of:

- The Pre-Turn Phase: Three steps before turn initiation (Steps-3 to-1).
- The Turn Event: The turning phase itself (Step 0).
- The Recovery Phase: Five steps immediately following turn completion (Steps +1 to +5).

The instantaneous arm-leg Phase Locking Value (PLV) was calculated for each step within this window, then averaged across a participant’s turns to give one trajectory per participant per side. Participants whose recordings never provided three clean pre-turn steps and five clean post-turn steps were excluded. The pre-turn steps ( 3 to 1) provide the pre-perturbation baseline shown in Figure 5a and were compared between groups separately; the recovery rate was defined as the linear slope of the PLV trajectory from the Turn Event (Step 0) to the third post-turn step (Step +3), capturing the acute phase of recovery.

Differences in recovery dynamics between frailty groups were assessed using a Linear Mixed-Effects Model (LMM) to account for repeated measures (left and right sides within a participant, and repeated steps within a trajectory) and subject-specific baselines. The model was specified as:

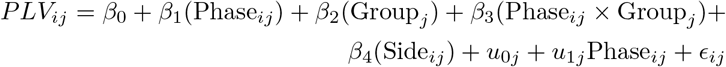

where Phase*_ij_* represents the step index relative to the turn (0, 1, 2, 3), Group*_j_* is the frailty status (Non-frail, Pre-frail, Frail) based on the Fried Frailty Phenotype [48], and Side*_ij_* distinguishes the two contralateral arm–leg pairs. The random effects *u*_0_*_j_* and *u*_1_*_j_* are the subject-specific intercept and slope for Phase. A random slope for Phase is required because the hypothesis concerns a group difference in slope; both it (*χ*^2^(2) = 393.6) and the Side term (*χ*^2^(1) = 30.2) were supported by likelihood-ratio test (both *p <* 0.001). Each participant was weighted by their turn count, since a trajectory averaged over more turns is more precisely estimated. Sensitivity analyses without weights, with turn count as a covariate and as an interaction with step, and with the sample restricted at turn-count thresholds of three, five and ten are reported in Table A4. In this model, the fixed effect coefficients were interpreted as follows:

- *â*_1_ (Main Effect of Phase): Represents the baseline recovery rate (slope) for the healthy (Non-frail) reference group.
- *â*_3_ (Interaction Effect): Represents the difference in recovery slope between the Frail group and the Healthy group. A negative *β*_3_ indicates a slower recovery.

To quantify the magnitude of the deficit, the percentage reduction in recovery rate was calculated as:

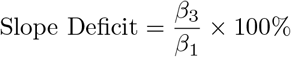

This metric isolates the relative loss of dynamic resilience in frail individuals compared to the healthy physiological standard.

To ensure that our findings were not dependent on the specific length of the recovery window, we repeated the LMM analysis across varying window lengths (Steps +2, +3, +4, and +5), keeping the same random-effects structure. The interaction effect (*β* slope) was evaluated for stability of direction and for statistical significance across all windows.

#### 5.3.5 Modelling of arm swing across age

To describe how the extracted upper-limb parameters vary with age, we employed Generalised Additive Models for Location, Scale and Shape (GAMLSS) [49]. The GAMLSS framework allows for the flexible modeling of a distribution’s mean, variance, and skewness as smooth functions of chronological age. Because the statistical properties of our metrics differ, distribution families were selected accordingly. For Arm Swing Amplitude, models were fitted using the Box-Cox Cole and Green (BCCG) distribution, which handles strictly positive, continuous data and accounts for potential age-dependent skewness in kinematic amplitudes. Conversely, because the arm-leg PLV is a coordination index strictly bounded between the intervals of [0, 1], we used a Beta Inflated (BEINF) distribution to accurately model its boundaries and variance. Following model optimization, continuous centile curves (i.e., 5th, 25th, 50th, 75th and 95th percentiles) were extracted to visualise the physiological aging trajectories of both locomotor capacity and neural coordination (performed using the gamlss package in R, version 5.5-0 [50]).

#### 5.3.6 Comparison of Arm Swing between Parkinson’s Disease Patients and Healthy Controls

Arm swing parameters were compared between groups at two levels. At the *participant level*, segmented data were averaged per participant, and group differences were tested using independent-samples *t*-tests with Hedges’ *g* as the effect size. At the *segment level*, a linear mixed-effects model (LMM) was fit for each outcome (Outcome ~ 1 + Group + (1 Participant ID)), accounting for within-participant segment correlation. The fixed-effect estimate is reported in the original units of each measure in Table 4; dividing it by *σ*^2^ + *σ*^2^ gives a standardised effect size comparable to Hedges’ *g*. The participant level analysis is treated as the inferential results and provide effect sizes for the two groups; the LMM results are reported as supporting evidence.

Statistical significance was set at *α* = 0.05 (two-tailed) throughout. Signal processing, parameter extraction, validation analyses and the Parkinson’s disease comparison were carried out in MATLAB R2025b [38]; the turn-recovery mixed models and the GAMLSS age models were fitted in R (lme4 and gamlss). No correction for multiple comparisons was applied. The analyses are organised as a small number of pre-specified questions rather than a screen, and each is reported with its effect size and confidence interval; the exception is the three-outcome Parkinson’s disease comparison, where a Bonferroni-corrected threshold of *α* = 0.017 would leave all three results significant. The exploratory factor analyses are descriptive and are not accompanied by hypothesis tests.

## Supplementary information

## Acknowledgements

The research was conducted at the Future Health Technologies at the Singapore-ETH Centre, which was established collaboratively between ETH Zürich and the National Research Foundation Singapore. This research is supported by the National Research Foundation Singapore (NRF) under its Campus for Research Excellence and Technological Enterprise (CREATE) programme. The data collection for the lab-based treadmill cohort and the corresponding research positions was funded by Innosuisse (Swiss Innovation Agency), Grant Number 120.497 IP-LS.

## Author Contributions

K.T. conceptualised the work. The study design and data acquisition were conducted by sub-teams specific to each cohort: Y.K, A.F., M.G., W.T., N.S. for the lab-based study; K.Z.T., K.Y.T., V.K., W.T., R.M., A.C., D.M. and N.S. for the overground home-based study; D.R., M.G., and N.S. for the Parkinson’s Disease study. K.T. prepared the initial draft of the manuscript. All authors actively participated in the comprehensive review, refinement, and finalization of the manuscript. N.S. supervised the project. All authors have read and agreed to the published version of the manuscript.

## Declarations

The authors declare no competing interests.

## Data availability

The wearable sensor gait data from both the lab-based treadmill cohort and the homebased overground walking cohort are not publicly available. Requests for access to the dataset can be directed to.

## Code availability

The arm swing algorithm and analysis code were implemented in MATLAB R2025b and R, and are available under the MIT license in the following GitLab repository: https://github.com/kaizedi/arm-swing-imu.

## Appendix A Supplementary materials

**Table A1:** Comparison of kinematic tracking accuracy between the Proposed Method and existing integration-based algorithms (Fan et al., Warmerdam et al.). Values are presented as Mean (Standard Deviation).

| Metric | Fan et al. | Warmerdam et al. | Proposed Method |
| --- | --- | --- | --- |
| RMSE ($^{\circ}$) | 8.3 (4.9) | 8.8 (4.1) | **3.6 (2.0)** |
| Pearson's $r$ | 0.77 (0.23) | 0.77 (0.38) | **0.94 (0.17)** |
| CMC | 0.70 (0.24) | 0.74 (0.23) | **0.93 (0.17)** |

RMSE: Root Mean Square Error; CMC: Coefficient of Multiple Correlation.

**Table A2:** Validation of gait event detection performance against the optical motion capture gold standard. Mean Error represents the temporal difference (ms) between IMU and Vicon events.

| Event | Mean Error (ms)<br>(SD) | Sensitivity<br>(%) | PPV<br>(%) | F1 Score |
| --- | --- | --- | --- | --- |
| Max Forward Swing (FS) | -2.4 (8.6) | 99.5 (1.0) | 99.2 (2.7) | 0.99 (0.02) |
| Max Backward Swing (BS) | 6.3 (14.4) | 99.3 (1.6) | 99.0 (2.9) | 0.99 (0.02) |

PPV: Positive Predictive Value; SD: Standard Deviation.

**Table A3:** Agreement between Wearable (IMU) and Optical (Ref) systems for derived gait parameters. Values indicate Mean (Standard Deviation) unless otherwise noted.

| Parameter | Ref Mean (SD) | IMU Mean (SD) | Bias (units) | Mean Abs. Error (%) | ICC (A,1) | LoA Width |
| --- | --- | --- | --- | --- | --- | --- |
| Arm Swing Time (s) | 1.14 (0.09) | 1.14 (0.09) | 0.000 s | 0.20% | 0.998 | 0.023 s |
| Arm Swing Amplitude (°) | 43.15 (15.20) | 39.66 (14.04) | -3.491° | 9.20% | 0.936 | 15.863° |
| Variability - Time (%CV) | 1.86 (0.93) | 1.97 (0.92) | 0.110 %CV | 9.90% | 0.966 | 0.859 %CV |
| Variability - Amplitude (%CV) | 10.97 (5.21) | 11.64 (4.99) | 0.672 %CV | 10.80% | 0.963 | 4.803 %CV |
| Asymmetry (Amplitude %) | 7.66 (6.23) | 8.00 (6.24) | 2.554 (MAE)* | 33.30% | 0.848 | 13.496 % |
| Coordination (PLV) | 0.92 (0.03) | 0.92 (0.04) | -0.004 | 1.10% | 0.929 | 0.052 |

\*For Asymmetry, the Bias column reports Mean Absolute Error (MAE). Bias is expressed in the units of the parameter; Mean Abs. Error is expressed relative to the reference mean. ICC: Intraclass Correlation Coefficient, single-measure absolute-agreement form; LoA: Limits of Agreement (95%), reported as the full width of the interval. Statistics are computed on participant means with the two sides averaged.

**Table A4:**
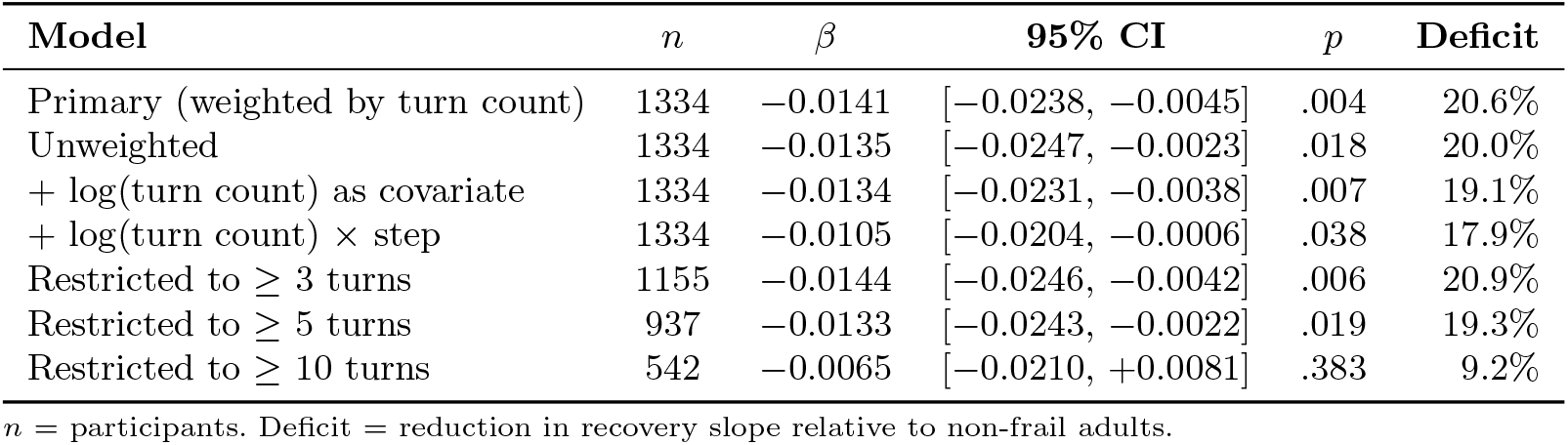
Sensitivity of the turn-recovery result to the handling of turn count. Each row reports the frail *×* step interaction, the difference in recovery slope between frail and non-frail adults, from the model described in the first column. The primary model weights each participant by the number of turns contributing to their trajectory.

## References

[1] Collins, S. H., Adamczyk, P. G. & Kuo, A. D. Dynamic arm swinging in human walking. Proceedings of the Royal Society B: Biological Sciences 276, 3679–3688 (2009). URL 10.1098/rspb.2009.0664.

[2] Meyns, P., Bruijn, S. M. & Duysens, J. The how and why of arm swing during human walking. Gait & Posture 38, 555–562 (2013). URL 10.1016/j.gaitpost.2013.02.006.

[3] Dean, J. C., Alexander, N. B. & Kuo, A. D. The effect of lateral stabilization on walking in young and old adults. IEEE Transactions on Biomedical Engineering 54, 1919–1926 (2007). URL 10.1109/TBME.2007.901031.

[4] Bruijn, S. M., Meijer, O. G., Beek, P. J. & van Dieën, J. H. The effects of arm swing on human gait stability. Journal of Experimental Biology 213, 3945–3952 (2010). URL 10.1242/jeb.045112.

[5] Wu, Y. et al. Effect of active arm swing to local dynamic stability during walking. Human Movement Science 45, 102–109 (2016). URL 10.1016/j.humov.2015.10.005.

[6] Ogata, T., Wen, B., Ye, R. & Miyake, Y. Gait training of healthy older adults in a sitting position using the wearable robot to assist arm-swing rhythm, walk-mate robot. Scientific Reports 14 (2024). URL 10.1038/s41598-024-76676-4.

[7] Gerards, M. H., McCrum, C., Mansfield, A. & Meijer, K. Perturbation-based balance training for falls reduction among older adults: Current evidence and implications for clinical practice. Geriatrics & Gerontology International 17, 2294–2303 (2017). URL 10.1111/ggi.13082.

[8] Shah, V. V. et al. Laboratory versus daily life gait characteristics in patients with multiple sclerosis, parkinson’s disease, and matched controls. Journal of NeuroEngineering and Rehabilitation 17 (2020). URL 10.1186/s12984-020-00781-4.

[9] Del Din, S. et al. Analysis of free-living gait in older adults with and without parkinson’s disease and with and without a history of falls: Identifying generic and disease-specific characteristics. The Journals of Gerontology: Series A 74, 500–506 (2017). URL 10.1093/gerona/glx254.

[10] Fan, B. et al. Imu-based real-time biofeedback wristband with automatic sensor-to-segment calibration for arm swing training. IEEE Sensors Journal 25, 9780–9789 (2025). URL 10.1109/JSEN.2025.3529416.

[11] Warmerdam, E. et al. Quantification of arm swing during walking in healthy adults and parkinson’s disease patients: Wearable sensor-based algorithm development and validation. Sensors 20, 5963 (2020). URL 10.3390/s20205963.

[12] Mirelman, A., et al. Effects of aging on arm swing during gait: The role of gait speed and dual tasking. PLOS ONE 10, e0136043 (2015). URL 10.1371/journal.pone.0136043.

[13] Ferraris, C. et al. Evaluation of arm swing features and asymmetry during gait in parkinson’s disease using the azure kinect sensor. Sensors 22, 6282 (2022). URL 10.3390/s22166282.

[14] Mirelman, A. et al. Arm swing as a potential new prodromal marker of parkinson’s disease. Movement Disorders 31, 1527–1534 (2016). URL 10.1002/mds.26720.

[15] Huang, X. et al. Both coordination and symmetry of arm swing are reduced in parkinson’s disease. Gait & Posture 35, 373–377 (2012). URL 10.1016/j.gaitpost.2011.10.180.

[16] Karatsidis, A. et al. Characterizing gait in people with multiple sclerosis using digital data from smartphone sensors: A proposed framework. Multiple Sclerosis Journal 31, 512–528 (2025). URL 10.1177/13524585251316242.

[17] Lee-Confer, J. Strength in arms: Empowering older adults against the risk of slipping and falling (2023). URL 10.51224/SRXIV.361.

[18] Leach, J. M., Mellone, S., Palumbo, P., Bandinelli, S. & Chiari, L. Natural turn measures predict recurrent falls in community-dwelling older adults: a longitudinal cohort study. Scientific Reports 8 (2018). URL 10.1038/s41598-018-22492-6.

[19] Thigpen, M. T., Light, K. E., Creel, G. L. & Flynn, S. M. Turning difficulty characteristics of adults aged 65 years or older. Physical Therapy 80, 1174–1187 (2000). URL 10.1093/ptj/80.12.1174.

[20] Mei, Z. et al. The role of electrode placement in subthalamic nucleus deep brain stimulation for improving gait in parkinson’s disease. Clinical Neurophysiology 182, 2111468 (2026). URL 10.1016/j.clinph.2025.2111468.

[21] Amprimo, G., Mei, Z., Ferraris, C., Olmo, G. & Ravi, D. A data-driven exploration and prediction of deep brain stimulation effects on gait in parkinson’s disease. IEEE Journal of Biomedical and Health Informatics 29, 4647–4658 (2025). URL 10.1109/jbhi.2024.3446548.

[22] Krasovsky, T. et al. Stability of gait and interlimb coordination in older adults. Journal of Neurophysiology 107, 2560–2569 (2012). URL 10.1152/jn.00950.2011.

[23] Hamacher, D., Singh, N., Van Dieën, J., Heller, M. & Taylor, W. Kinematic measures for assessing gait stability in elderly individuals: a systematic review. Journal of The Royal Society Interface 8, 1682–1698 (2011).

[24] Clark, D. J. Automaticity of walking: functional significance, mechanisms, measurement and rehabilitation strategies. Frontiers in Human Neuroscience 9 (2015). URL 10.3389/fnhum.2015.00246.

[25] Frigon, A. The neural control of interlimb coordination during mammalian locomotion. Journal of Neurophysiology 117, 2224–2241 (2017). URL 10.1152/jn.00978.2016.

[26] Swinnen, S. P. Intermanual coordination: From behavioural principles to neuralnetwork interactions. Nature Reviews Neuroscience 3, 348–359 (2002). URL 10.1038/nrn807.

[27] Middleton, F. Basal ganglia and cerebellar loops: motor and cognitive circuits. Brain Research Reviews 31, 236–250 (2000). URL 10.1016/S0165-0173(99)00040-5.

[28] Blandini, F., Nappi, G., Tassorelli, C. & Martignoni, E. Functional changes of the basal ganglia circuitry in parkinson’s disease. Progress in Neurobiology 62, 63–88 (2000). URL 10.1016/s0301-0082(99)00067-2.

[29] Stuart, S., Belluscio, V., Quinn, J. F. & Mancini, M. Pre-frontal cortical activity during walking and turning is reliable and differentiates across young, older adults and people with parkinson’s disease. Frontiers in Neurology 10 (2019). URL 10.3389/fneur.2019.00536.

[30] Montero-Odasso, M., Verghese, J., Beauchet, O. & Hausdorff, J. M. Gait and cognition: A complementary approach to understanding brain function and the risk of falling. Journal of the American Geriatrics Society 60, 2127–2136 (2012). URL 10.1111/j.1532-5415.2012.04209.x.

[31] Seidler, R. D. et al. Motor control and aging: Links to age-related brain structural, functional, and biochemical effects. Neuroscience & Biobehavioral Reviews 34, 721–733 (2010). URL 10.1016/j.neubiorev.2009.10.005.

[32] Maki, B. E. Gait changes in older adults: Predictors of falls or indicators of fear? Journal of the American Geriatrics Society 45, 313–320 (1997). URL 10.1111/j.1532-5415.1997.tb00946.x.

[33] Renggli, D. et al. Wearable inertial measurement units for assessing gait in realworld environments. Frontiers in Physiology 11 (2020).

[34] Kim, Y. K. et al. Leveraging deep learning and wearables for automatically identifying gait event: Effects of age and location of sensors on the assessment of gait events. IEEE Sensors Journal 25, 792–802 (2025).

[35] Ho, D. E., Imai, K., King, G. & Stuart, E. A. ¡b¿matchit¡/b¿: Nonparametric preprocessing for parametric causal inference. Journal of Statistical Software 42 (2011). URL 10.18637/jss.v042.i08.

[36] Laidig, D. & Seel, T. Vqf: Highly accurate IMU orientation estimation with bias estimation and magnetic disturbance rejection. Information Fusion 91, 187–204 (2023).

[37] Tan, K. Z. et al. Walking in the free world: Establishing normative trajectories for ecological assessment of robust gait variability with age (2026). URL 10.64898/2026.03.06.26347806.

[38] Inc., T. M. Signal processing toolbox version r2025b (2022). URL https://www.mathworks.com.

[39] Zifchock, R. A., Davis, I., Higginson, J. & Royer, T. The symmetry angle: A novel, robust method of quantifying asymmetry. Gait & Posture 27, 622–627 (2008). URL 10.1016/j.gaitpost.2007.08.006.

[40] Plotnik, M., Giladi, N. & Hausdorff, J. M. A new measure for quantifying the bilateral coordination of human gait: effects of aging and parkinson’s disease. Experimental Brain Research 181, 561–570 (2007). URL 10.1007/s00221-007-0955-7.

[41] Lamb, P. F. & Stöckl, M. On the use of continuous relative phase: Review of current approaches and outline for a new standard. Clinical Biomechanics 29, 484–493 (2014). URL 10.1016/j.clinbiomech.2014.03.008.

[42] Aydore, S., Pantazis, D. & Leahy, R. M. A note on the phase locking value and its properties. NeuroImage 74, 231–244 (2013). URL 10.1016/j.neuroimage.2013.02.008.

[43] Kadaba, M. P. et al. Repeatability of kinematic, kinetic, and electromyographic data in normal adult gait. Journal of Orthopaedic Research 7, 849–860 (1989). URL 10.1002/jor.1100070611.

[44] Koo, T. K. & Li, M. Y. A guideline of selecting and reporting intraclass correlation coefficients for reliability research. Journal of Chiropractic Medicine 15, 155–163 (2016).

[45] Martin Bland, J. & Altman, D. Statistical methods for assessing agreement between two methods of clinical measurement. The Lancet 327, 307–310 (1986). URL 10.1016/S0140-6736(86)90837-8.

[46] Bustos Navarrete, C. & Coutinho Soares, F. dominanceanalysis: Dominance analysis (2019). URL 10.32614/CRAN.package.dominanceanalysis.

[47] Revelle, W. *psych: Procedures for Psychological, Psychometric, and Personality Research*. Northwestern University, Evanston, Illinois (2025). URL https://CRAN.R-project.org/package=psych. R package version 2.5.6.

[48] Fried, L. P. et al. Frailty in older adults: Evidence for a phenotype. The Journals of Gerontology Series A: Biological Sciences and Medical Sciences 56, M146–M157 (2001). URL 10.1093/gerona/56.3.m146.

[49] Rigby, R. A., Stasinopoulos, M. D., Heller, G. Z. & De Bastiani, F. Distributions for modeling location, scale, and shape: Using gamlss in r (2019). URL 10.1201/9780429298547.

[50] Rigby, R. A. & Stasinopoulos, D. M. Generalized additive models for location, scale and shape. Journal of the Royal Statistical Society Series C: Applied Statistics 54, 507–554 (2005). URL 10.1111/j.1467-9876.2005.00510.x.

